# Disruption of RFX family transcription factors causes autism, attention deficit/hyperactivity disorder, intellectual disability, and dysregulated behavior

**DOI:** 10.1101/2020.09.09.20187104

**Authors:** Holly K. Harris, Tojo Nakayama, Jenny Lai, Boxun Zhao, Nikoleta Argyrou, Cynthia S. Gubbels, Aubrie Soucy, Casie A. Genetti, Lance H. Rodan, George E. Tiller, Gaetan Lesca, Karen W. Gripp, Reza Asadollahi, Ada Hamosh, Carolyn D. Applegate, Peter D. Turnpenny, Marleen E.H. Simon, Catharina (Nienke) M.L. Volker-Touw, Koen L.I. van Gassen, Ellen van Binsbergen, Rolph Pfundt, Thatjana Gardeitchik, Bert B.A. de Vries, Ladonna L. Imken, Catherine Buchanan, Marcia Willing, Tomi L. Toler, Emily Fassi, Laura Baker, Fleur Vansenne, Xiadong Wang, Julian L. Ambrus, Madeleine Fannemel, Jennifer E. Posey, Emanuele Agolini, Antonio Novelli, Anita Rauch, Paranchai Boonsawat, Christina R. Fagerberg, Martin J. Larsen, Maria Kibaek, Audrey Labalme, Alice Poisson, Katelyn K. Payne, Laurence E. Walsh, Kimberly Aldinger, Jorune Balciuniene, Cara Skraban, Christopher Gray, Jill Murrell, Caleb P. Bupp, Giulia Pascolini, Paola Grammatico, Martin Broly, Sébastien Küry, Mathilde Nizon, Iqra Ghulam Rasool, Muhammad Yasir Zahoor, Cornelia Kraus, André Reis, Muhammad Iqbal, Kevin Uguen, Severine Audebert-Bellanger, Claude Ferec, Sylvia Redon, Janice Baker, Yunhong Wu, Guiseppe Zampino, Steffan Syrbe, Ines Brosse, Rami Abou Jamra, William B. Dobyns, Lilian L. Cohen, Pankaj B. Agrawal, Alan Beggs, Timothy W. Yu

**Author notes:** These authors contributed equally to this work (Holly K. Harris, Tojo Nakayama). **Correspondence to:** Timothy W. Yu, Division of Genetics and Genomics, Boston Children’s Hospital, 300 Longwood Avenue, Mailstop BCH3150, Boston, MA 02115.

## Abstract

**Purpose:** We describe a novel neurobehavioral syndrome of autism spectrum disorder, intellectual disability, and attention deficit/hyperactivity disorder associated with *de novo* or inherited deleterious variants in members of the *RFX* family of genes. *RFX* genes are evolutionarily conserved transcription factors that act as master regulators of central nervous system development and ciliogenesis.

**Methods:** We assembled a cohort of 36 individuals (from 31 unrelated families) with *de novo* mutations in *RFX3, RFX4*, and *RFX7*. We describe their common clinical phenotypes and present bioinformatic analyses of expression patterns and downstream targets of these genes as they relate to other neurodevelopmental risk genes.

**Results:** These individuals share neurobehavioral features including autism spectrum disorder (ASD), intellectual disability, and attention-deficit/hyperactivity disorder (ADHD); other frequent features include hypersensitivity to sensory stimuli and sleep problems. *RFX3, RFX4*, and *RFX7* are strongly expressed in developing and adult human brain, and X-box binding motifs as well as *RFX* ChIP-seq peaks are enriched in the cis-regulatory regions of known ASD risk genes.

**Conclusion:** These results establish deleterious variation in *RFX3, RFX4*, and *RFX7* as important causes of monogenic intellectual disability, ADHD and ASD, and position these genes as potentially critical transcriptional regulators of neurobiological pathways associated with neurodevelopmental disease pathogenesis.

## INTRODUCTION

Autism spectrum disorder (ASD), marked by deficits in social communication and the presence of restricted interests and repetitive behavior,^1^ is highly heritable and genetically heterogeneous,^2^ with *de novo* loss-of-function variants known to be an important contributor to ASD risk.^3–5^ ASD is often comorbid with other neurodevelopmental diagnoses, including attention-deficit/hyperactivity disorder (ADHD), a behavioral diagnosis that captures a persistent pattern of inattention and/or hyperactivity-impulsivity that interferes with functioning.^1^ Emerging evidence also points to a role of *de novo* loss-of-function variants in ADHD,^6–8^ and recent analyses of genome-wide association studies from over 20,000 individuals with ADHD suggests a role for common variation at 12 significant risk loci, implicating genes involved in neurotransmitter regulation and neuronal plasticity.^9^

*RFX3* is a member of the regulatory factor X (*RFX*) gene family which encodes transcription factors with a highly-conserved DNA binding domain. Located on chromosome 9p24.2, the gene is comprised of 307,708 base pairs and encodes a transcribed protein that is 749 amino acids in length. *RFX3* is expressed in several tissues including developing and adult brain. Other *RFX* family members *(RFX1, 4, 5*, and 7) are also highly expressed in brain tissue^10–13^, with expression patterns of *RFX1, 3, 4* and *7* clustering especially tightly.^13^

We report a series of 36 individuals from 31 families with deleterious, mostly *de novo* variants in three brain-expressed members of the *RFX* family: *RFX3, RFX4*, or *RFX7. RFX3* was among 102 genes recently identified as statistically enriched for *de novo* variants in a large-scale analysis of trio exome data from individuals with ASD,^14^ but to date *RFX4* and *RFX7* have not been previously associated with human disease. Analysis of their clinical presentations reveals common features including intellectual disability (ID), ASD, and/or ADHD, delineating a novel neurobehavioral phenotype associated with *RFX* haploinsufficiency.

## MATERIALS AND METHODS

### Ethics Statement

This series was compiled via an international collaborative effort involving Boston Children’s Hospital, Kaiser Permanente, Lyon University Hospital, Nemours/A.I. DuPont Hospital for Children, University of Zurich, Johns Hopkins University, Peninsula Clinical Genetics at Royal Devon and Exeter NHS Foundation Trust, University Medical Centre Utrecht, Radboud University Medical Centre, Dell Children’s Medical Group, Washington University School of Medicine in St. Louis, University Medical Center Groningen, Ciphergene, SUNY at Buffalo School of Medicine, Oslo University Hospital, Baylor College of Medicine, Bambino Gesu Children’s Hospital, Odense University Hospital, Indiana University Health Neuroscience Center, Seattle Children’s Research Institute, Children’s Hospital of Philadelphia, Spectrum Health Helen DeVos Children’s Hospital, Sapienza University and San Camillo-Forlanini Hospital, CHU Nantes et Service de Génétique Médicale, University of Erlangen-Nuremberg, The Islamia University of Bahawalpur, Brest University Hospital, Children’s Minnesota, Universita Cattolica del Sacro Cuore, University Hospital Heidelberg, University of Leipzig Medical Center, University of Washington, and Weill Cornell Medical College. Collaboration was facilitated by the online genetics/genomics resource GeneMatcher. Affected individuals were clinically assessed by at least one clinical geneticist from one of the participating centers. De-identified clinical data from collaborating institutions (collected with local IRB approval or deemed exempt from IRB review as per local institutional policy) was shared for analysis and publication under a study protocol approved by the Boston Children’s Hospital IRB.

### Case Ascertainment and Data Collection

We obtained phenotypic data from 15 unrelated individuals with loss-of-function variants in *RFX3*, 3 unrelated individuals with loss-of-function variants in *RFX4*, and 13 unrelated individuals with loss-of-function variants in *RFX7*. De-identified individual case summaries are available upon request. Variants arose *de novo* with the exception of four related individuals from the same nuclear family with the same heterozygous loss-of-function variant in *RFX3*, and three other related cases in *RFX4* (homozygous for an inherited missense variant). Pedigree information is shown in Figure 1. Diagnoses of ASD were reported in the medical record, but not uniformly evaluated by standardized measures such as the Diagnostic and Statistical Manual, Fourth or Fifth Edition (DSM-IV and 5),^1,15^ Autism Diagnostic Observation Schedule (ADOS),^16^ or Autism Diagnostic Interview, Revised (ADI-R).^17^ Similarly, ID and ADHD diagnoses were accepted per clinician report and not always accompanied by standardized cognitive or behavioral testing measures.

**Figure 1.**
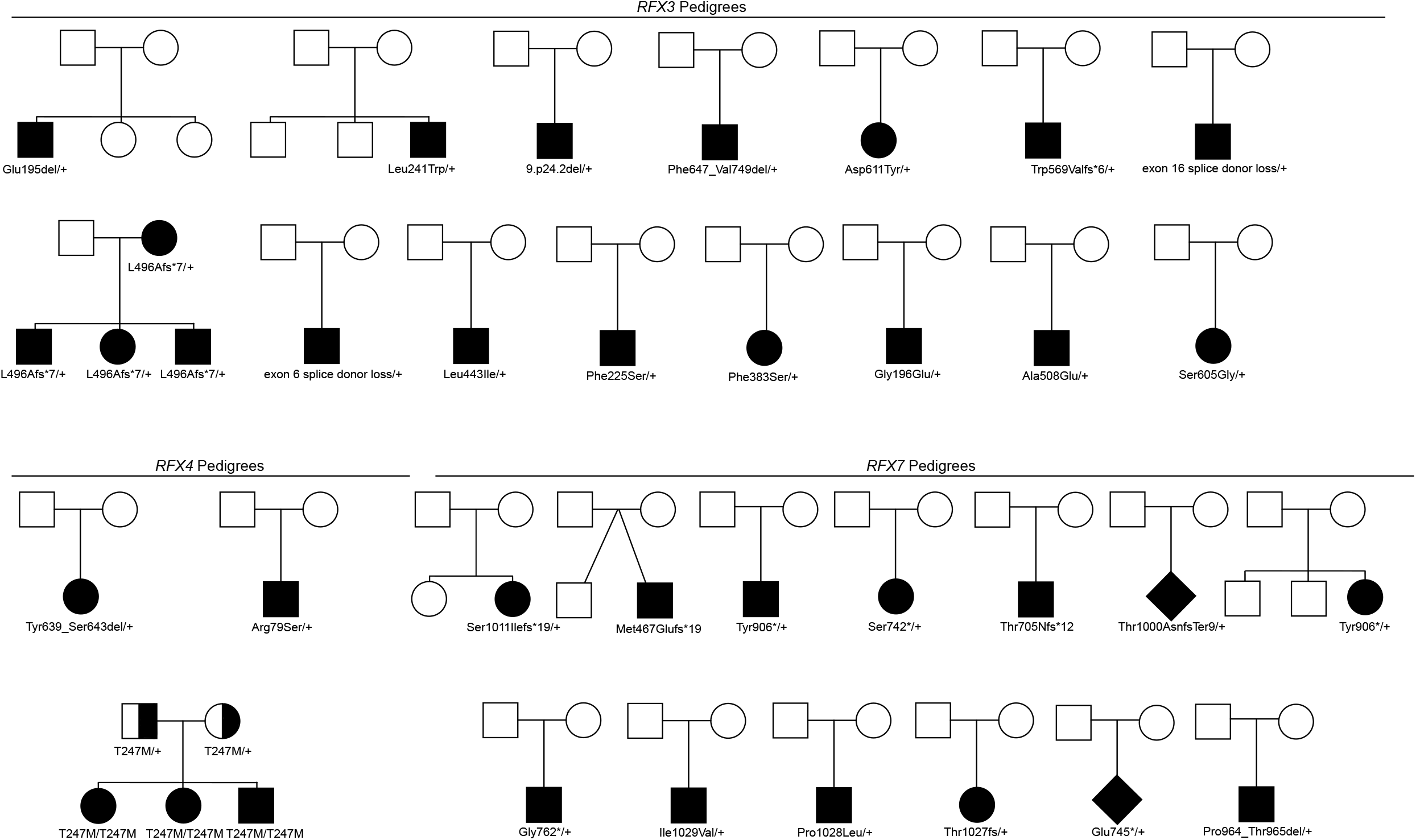
Individuals with Pathogenic Variants in *RFX* Members. *RFX3, RFX4*, and *RFX7* case pedigrees. All mutations were de novo except for p.(Leu496Alafs*7) which was found in an affected mother and three children, and p.(Thr247Met), found in the homozygous state in three affected children.

### Exome Sequencing

Individuals included underwent exome sequencing on a clinical or research basis. Seven of the individuals were sequenced through GeneDx using genomic DNA from the proband or proband plus parents, captured using either the Clinical Research Exome kit (Agilent Technologies, Santa Clara, CA) or the IDT xGen Exome Research Panel v1.0, and sequenced on an Illumina system with 100bp or greater paired-end reads. Reads were aligned to human genome build GRCh37/UCSC hg19, and variants were analyzed and interpreted as previously described^18^ using variant classification criteria publicly available on the GeneDx ClinVar submission page (see Web Resources). Two cases of *RFX7* were sequenced through Ambry Genetics whose gene and variant classification process are available on the AmbryGenetics web page. The remainder of the individual’s exome sequencing was performed through the clinicians’ institutions or an external laboratory or research program (see Acknowledgments).

### Variant Analyses

Variant genomic coordinates are reported in relation to the Human Dec. 2013 (GRCh38/hg38) Assembly. The reference mRNA and protein sequences used are *RFX3* NM_134428.2, NP_602304.1; *RFX4* NM_213594.2, NP_998759.1; and *RFX7*NM_022841.5, NP_073752.5. The variant databases gnomAD v2.1.1 and v3 were examined for the presence of each variant.^19^ Predicted pathogenicities for all variants were assessed by MutationTaster^20^ (Schwarz et al., 2010). For missense variants, pathogenicity predictions were obtained from six algorithms: MutationTaster,^20^ SIFT,^21^ PolyPhen2,^22^ PROVEAN,^23^ LRT,^24^ and MutationAssessor,^25^ and the total number of algorithms out of six with a deleterious prediction is referred to as the Nonsynonymous Damaging score (NsynD). In addition, the regional missense constraint metric MPC score^26^ and CADD scaled score^27^ are reported in Table S2 and Table S3. MPC ≥ 2 is considered highly deleterious.

### Cell transfection and culture

Human *RFX3* (NM_134428.2; Human *RFX3* cDNA) was cloned into V5-or Myc-tagged mammalian expression vectors using the Gateway cloning system (Thermo Fisher Scientific). Point mutations were introduced with the QuikChange Lighting Site-Directed Mutagenesis kit (Agilent Technologies) to incorporate variants from affected individuals. To quantify the expression level of exogenous *RFX3*, equal amounts of tagged-RFX3 expression vectors were transfected into Hela cells or HEK293T using Lipofectamine 3000 (Thermo Fisher Scientific). The transfected cells were cultured for 48 hours before harvesting. Cell extracts were analyzed by immunoblotting, using antibodies raised against RFX3 (HPA035689, Sigma-Aldrich), V5 (R960-25, Thermo Fisher Scientific), or beta actin (ab6276, Abcam). Blots were scanned on a Li-Cor Odyssey imager (Li-Cor). Signal intensities were quantified using Image Studio Lite (Li-Cor). Each immunoblot analysis was replicated three times.

### KEGG pathway and ASD gene set over-representation analysis

ChIP-seq and eCLIP-seq narrowPeak bed files for RFX family members, CREBBP, EP300, FMR1, FXR1, and FXR2 were obtained from the ENCODE portal,^28^ and additional ChIP-seq data for RFX3_K562 were obtained from RegulomeDB^29^ (Table S6). Functional binding genes (1 kb upstream/downstream of TSS [transcriptional start site]) were annotated using ChIPseeker.^30^ ASD risk gene lists included 102 TADA genes from Satterstrom *et al*. and 253 ASD/ID genes from Coe *et al*. (Table S7).^14,31^ Differentially expressed genes (DEGs) in ASD brains were extracted from Velmeshev *et al*. excluding endothelial DEGs that could originate from vascular cells in the brain (Table S7).^32^ Customized KEGG (Kyoto Encyclopedia of Genes and Genomes) pathway analysis was performed using clusterProfiler^33^ to determine the enrichment for KEGG pathways, ASD risk gene sets, and ASD DEGs. Multiple testing correction was performed using Benjamini-Hochberg correction (Table S8). Annotations, statistical analyses, and plots were implemented in R.

### Motif analysis

For motif occurrence analysis, FIMO was used to scan promoter sequences for individual occurrences of RFX motifs.^34^ For all analyses, motif models were obtained from the JASPAR 2020 database.^35^ Motifs searched for included RFX3 (MA0798.1), RFX4 (MA0799.1), and RFX7 (MA1554.1). Promoter sequences were defined as -1000 base pairs and +500 base pairs relative to the transcription start site. Motif occurrences were classified as significant based on a reporting threshold of p-value <0.00001 and q-value (Benjamini) <0.10. For motif enrichment analysis, we used the HOMER findMotifs.pl and findMotifsGenome.pl scripts.^36^ Motifs were classified as enriched based on fold-enrichment > 1.5 and q-value (Benjamini) <0.01. Enhancer sequences associated with genes of interest were obtained from Enhancer Atlas 2.0.^37^ ASD risk gene lists were obtained as noted previously to include 102 TADA genes and 124 ASD/ID genes reaching exome-wide significance.^14,31^

## RESULTS

**Index Case:** A teenaged male presented for genetic evaluation of ASD, ADHD, and borderline ID. His prenatal course was uncomplicated. Developmental delays in language, social communication, and motor functioning were first observed at 18 months of age. He had a history of sleep disturbance that improved by adolescence. He had notable deterioration of his behavioral functioning in adolescence, requiring placement in an intensive therapeutic program. His behavioral profile was marked by mood lability, anxiety, impulsivity, aggression, noise hypersensitivity and self-soothing motor stereotypies (e.g. rocking). An electroencephalogram (EEG) was obtained for a clinical concern for absence seizures but was normal. Medical history was otherwise unremarkable. Growth parameters were normal including head circumference. Physical examination was notable for subtle dysmorphisms including a high arched palate and down-turned corners of the mouth, mild hypotonia and generalized joint hyperlaxity. Strength, reflexes, and sensation were normal.

Exome sequencing revealed a heterozygous in-frame deletion of three nucleotides in the coding region of *RFX3* (c.584_586delGAA; p.(Glu195del)). This variant is absent from dbSNP and gnomAD and parental sequencing revealed the deletion to be *de novo*. It is predicted to lie within the conserved DNA binding domain of *RFX3* and removes an amino acid (glutamate-195) that is evolutionarily conserved in all 9/9 species examined down to zebrafish. *RFX3* is under significant loss-of-function constraint in databases of human population variation (pLI score 1.0 in ExAC, gnomAD.

### Case series of individuals with *RFX3* pathogenic variants

We identified and obtained clinical information from 18 individuals inclusive of our index case via GeneMatcher,^38^ all bearing loss-of-function variants in *RFX3*. Genotypic information is provided in Table 1, clinical phenotypes are summarized in Tables 2 and S1, and variant pathogenicity predictions are summarized in Supplemental Table S2. A total of 15 distinct variants were identified: two frameshift variants, two canonical splice donor variants, eight missense variants, one in-frame deletion, one 42 kb deletion removing the last two exons of *RFX3*, and one 227 kb deletion involving only *RFX3*. In one family, an affected parent transmitted a frameshift variant to three affected children; all other variants were *de novo* and novel (Figure 1).

**Table 1.** Molecular findings in individuals with ASD, ADHD, and/or ID and variants in *RFX3, RFX4*, or *RFX7*. Molecular characterization of *RFX3, RFX4*, and *RFX7* variants reported in this study. Chromosome structure is described according to the Human Dec. 2013 (GRCh38/hg38) Assembly. RefSeq identifiers: RFX3 NM_134428.2, NP_602304.1; RFX4 NM_213594.2, NP_998759.1; RFX7 NM_022841.5, NP_073752.5. Protein domains were obtained from Sugiaman-Trapman et al., 2018.^13^ Italics indicates individual has a variant of uncertain significance.

| Gene | Inheritance | gDNA (GRCh38) | cDNA | Protein | Category | Domain |
| --- | --- | --- | --- | --- | --- | --- |
| *RFX3* | *de novo* | chr9:g.3293222_3293224del | c.584_586del | p.Glu195del | Inframe deletion | DBD |
|  | *de novo* | chr9:g.3293086A>C | c.722T>G | p.Leu241Trp | Missense | DBD |
|  | *de novo* | chr9:g.9p24.2del | NA | (gene deletion) | Deletion | (all) |
|  | *de novo* | chr9:g.3197716_3239762del | c.1968+8270_*27326del | (exon 17 and exon 18 deleted) p.Phe647_Val749del | Deletion | DD |
|  | *de novo* | chr9:g.3248169C>A | c.1831G>T | p.Asp611Tyr | Missense | DD |
|  | *de novo* | chr9:g.3257101dupG | c.1704dupG | p.Trp569Valfs*6 | Frameshift | DD |
|  | *de novo* | chr9:g.3248031C>T | c.1968+1G>A | exon 16 splice donor loss | Splicing | DD |
|  | inherited | chr9:g.3263053_3263054del | c.1486_1487del | p.Leu496Alafs*7 | Frameshift | DD |
|  | *de novo* | chr9:g.3301541C>T | c.549+5G>A | exon 6 splice donor loss | Splicing | DBD |
|  | *de novo* | chr9:g.3270401G>T | c.1327C>A | p.Leu443Ile | Missense | EDD |
|  | *de novo* | chr9:g.3293134A>G | c.674T>C | p.Phe225Ser | Missense | DBD |
|  | *de novo* | chr9:g.3271057A>G | c.1148T>C | p.Phe383Ser | Missense | EDD |
|  | *de novo* | chr9:g.3293221C>T | c.587G>A | p.Gly196Glu | Missense | DBD |
|  | *de novo* | chr9:g.3263017G>T | c.1523C>A | p.Ala508Glu | Missense | DD |
|  | *de novo* | chr9:g.3256992T>C | c.1813A>G | p.Ser605Gly | Missense | DD |
| *RFX4* | *de novo* | chr12:g.106750773_106750787del | c.1915_1929del | p.Tyr639_Ser643del | Inframe deletion | NA |
|  | *de novo* | chr12:g.106654271C>A | c.235C>A | p.Arg79Ser | Missense | DBD |
|  | recessive (homozygous) | chr12:g.106696353C>T | c.740C>T | p.Thr247Met | Missense | NA |
| *RFX7* | *de novo* | chr15:g.56094696del | c.3032del | p.Ser1011Ilefs*19 | Frameshift | NA |
|  | unknown (adopted) | chr15:g.56096328_56096329del | c.1399_1400del | p.Met467Glufs*19 | Frameshift | NA |
|  | *de novo* | chr15:g.56095010G>T | c.2718C>A | p.Tyr906* | Stop Gain | NA |
|  | *de novo* | chr15:g.56095503G>C | c.2225C>G | p.Ser742* | Stop Gain | NA |
|  | *de novo* | chr15:g.56095615dupA | c.2113dupT | p.Thr705Nfs*12 | Frameshift | NA |
|  | *de novo* | chr15:g.56094730dupT | c.2998dupA | p.Thr1000AsnfsTer9 | Frameshift | NA |
|  | *de novo* | chr15:g.56095010G>C | c.2718C>G | p.Tyr906* | Stop Gain | NA |
|  | *de novo* | chr15:g.56095444C>A | c.2284G>T | p.Gly762* | Stop Gain | NA |
|  | *de novo* | chr15:g.56094643T>C | c.3085A>G | p.Ile1029Val | Missense | NA |
|  | *de novo* | chr15:g.56094645G>A | c.3083C>T | p.Pro1028Leu | Missense | NA |
|  | *de novo* | chr15:g.56094648del | c.3080del | p.Thr1027fs | Frameshift | NA |
|  | *de novo* | chr15:g.56095495C>A | c.2233G>T | p.Glu745* | Stop Gain | NA |
|  | *de novo* | chr15:g.56094864_56094869del | c.2859_2864del | p.Pro964_Thr965del | Inframe deletion | NA |

**Table 2.**
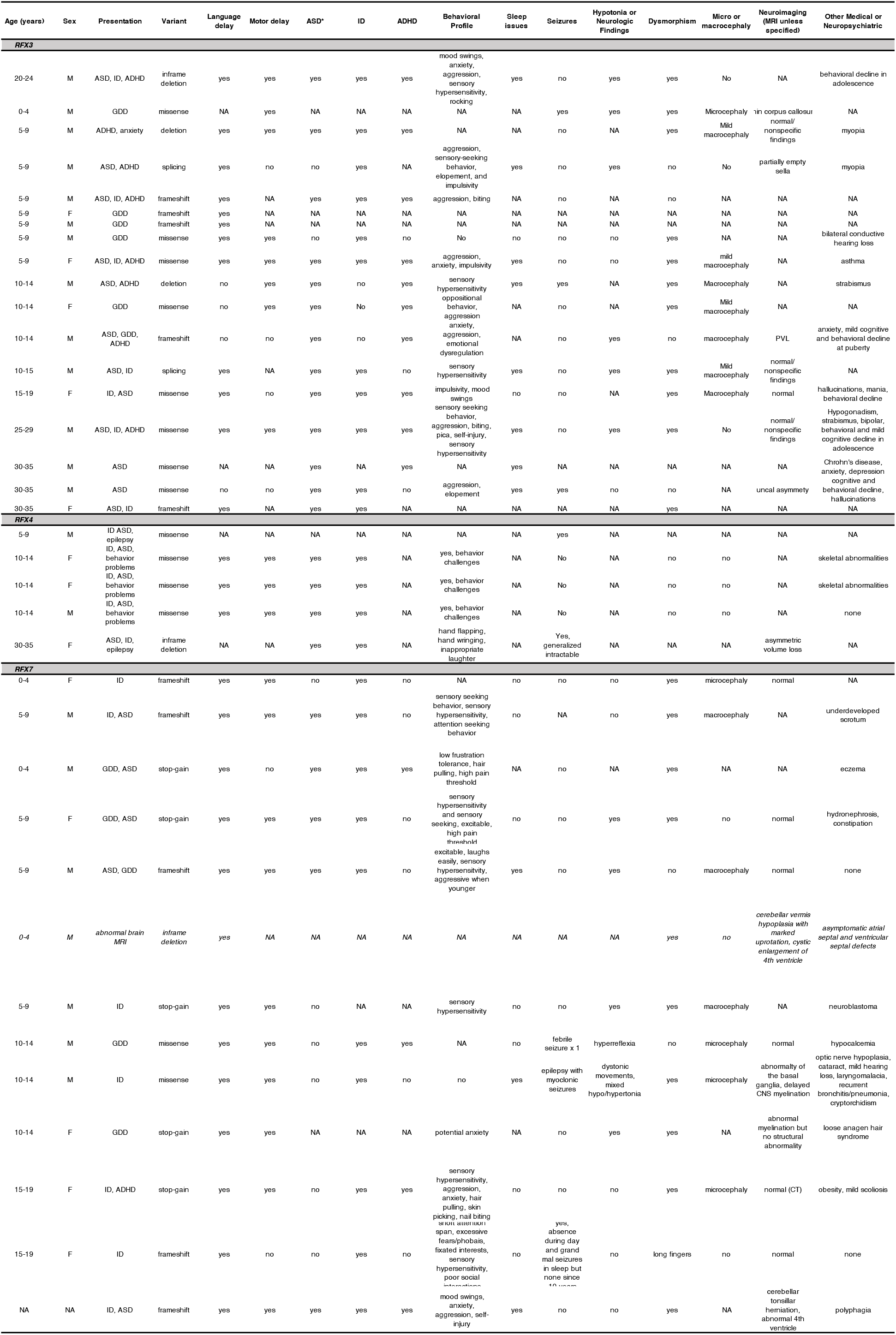
Clinical features of individuals with pathogenic variants in *RFX3, RFX4*, or *RFX7*. *ASD = We recognize the heterogeneity in ASD diagnoses. For our table, individuals were considered to have ASD if documented in the clinical note as having ASD diagnosed via formal measure, according to clinical expertise, or documented as having clear “autistic features,” a designation we considered equivalent to a diagnosis of ASD for purposes of this report. Granular description of social communication or restrictive and repetitive behavior data to determine DSM-5 diagnosis was not uniformly available. NA, not available. ID, intellectual disability. GDD, global developmental delay. ADHD, attention deficit hyperactivity disorder. MRI, magnetic resonance imaging. CT, computerized tomography. PVL, periventricular leukomalacia. Italics indicates individual has a variant of uncertain significance.

Phenotypic data from these 18 individuals (thirteen males and five females) are presented in Table 2. There were no sex-based differences in severity of phenotype. All individuals had neurodevelopmental delays, with formally recorded clinical diagnoses of ASD (72%) and ID (61%) of varying severity (borderline to moderate; described as global developmental delay in young children), and ADHD (56%) (Table S1). Many showed a distinct behavioral pattern marked by easy excitability/overstimulation, hypersensitivity to sensory (particularly auditory) stimuli, anxiety, emotional dysregulation and/or aggression (13/15 [87%] with specific behavioral information provided). Three individuals were reported to have seizures (17%). Some individuals had sleep difficulties (44%) including limited total duration of sleep, frequent awakenings, or early morning awakenings. Subtle non-specific and non-recurrent dysmorphisms were commonly reported (61%), including broad nasal bridge, high, arched palate, and hand and foot abnormalities (tapered fingers, widely spaced toes), but no consistent recognizable features were shared by all individuals. Both macrocephaly (six individuals) and microcephaly (two individuals) were reported (8/11 individuals [73%] with a head circumference measurement or percentile provided). Magnetic resonance imaging (MRI) of the brain was available for eight individuals, with reports of non-specific findings in four, including white matter changes, uncal asymmetry, partially empty sella, or prominent ventricles. One individual had mild thinning of the corpus callosum (Table 2). Five of seven individuals (71%) who were past the onset of puberty (ages 12-30 years) had reports of behavioral and/or cognitive worsening at the time of puberty/adolescence. Three had increased aggression specifically noted. Three were described as having manic and/or psychotic symptoms, specifically two described as having hallucinations (one requiring psychiatric hospitalization) and another described as having conversations with imaginary friends. Three were reported to have had decline in cognition, one in adolescence and another around 28 years of age.

### *RFX3* pathogenic variants lead to reduced RFX3 levels

The types of *RFX3* variants encountered strongly supported a loss-of-function model in which mutations disrupt or destabilize expression of RFX3 protein. We tested this hypothesis by engineering a subset of variants into *RFX3* cDNAs for expression analysis in Hela and HEK293 cells (Figure 2B). As expected, the frameshift variant (p.Trp569ValfsX6, predicted to cause nonsense-mediated decay of the mutant mRNA) led to essentially complete loss of RFX3 protein expression. Missense variants p.(Asp611Tyr), p.(Leu443Ile), p.(Phe383Ser), and the stop-gain variant p.(Glu195del) also led to significant decreases in RFX3 levels, suggesting that *RFX3* pathogenic variants lead to haploinsufficiency through reduced protein levels (Figure 2B).

**Figure 2.**
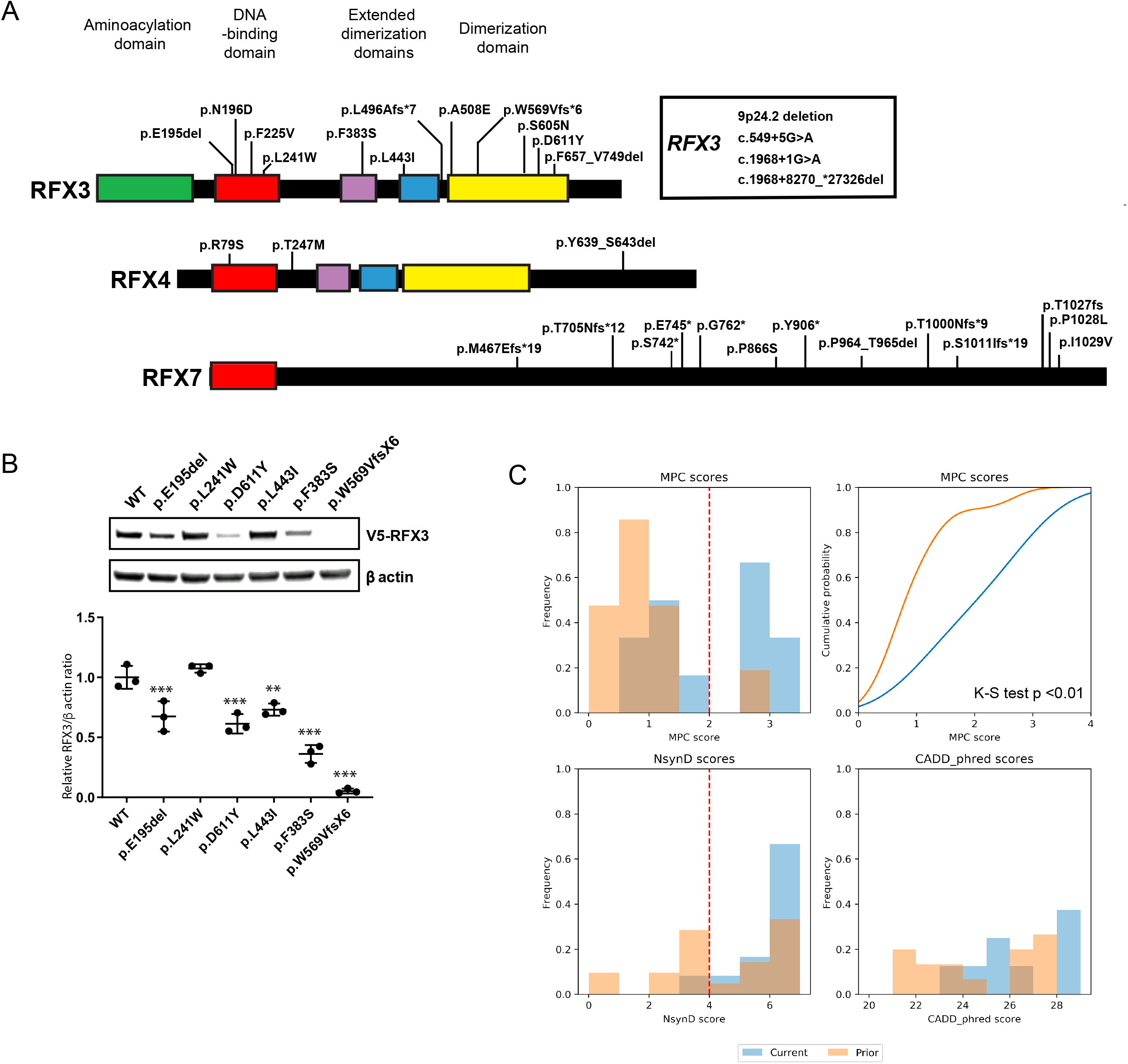
Characterization of *RFX* Variants. (A) Mapping of selected RFX variants to domains. Whole gene deletion and intronic variants are not illustrated. RFX3 (NP_602304.1), RFX4 (NP_998759.1), RFX7 (NP_073752.5). (B) Immunoblot analysis of RFX3 abundance in Hela cells overexpressed with RFX3 mutants. Quantification of RFX3 abundance was performed. Values represent the average of three independent experiments. Error bars represent standard deviation. ***p<0.001, **p<0.01. (C) Missense variant pathogenicity scores for the currently reported variants (current) and prior reported variants (prior) in *RFX3, 4*, and 7. The distribution of MPC scores for missense variants reported in this study is significantly different from that of prior reported missense variants, Kolmogorov-Smirnov (K-S) test p-value <0.01. MPC, Missense badness, PolyPhen-2, and Constraint. NsynD, Nonsynonymous Damaging score. CADD, Combined Annotation Dependent Depletion.

### Pathogenic variants in additional RFX family genes cause related neurodevelopmental phenotypes

Additional individuals were ascertained who harbored loss-of-fonction variants in other closely related genes of the *RFX* family. Thirteen individuals bearing *de novo* loss-of-fUnction variants in *RFX7* were identified (Tables 1 and 2), including three frameshift variants, five stop gain variants, one in-frame deletion, and two missense variants (Table 1, Table S2). Again, more males were identified than females (eight males, four females) without differences in phenotype based on sex. All individuals had language delay, and most had ID/global developmental delay (85%) (Tables 2 and 3). While formal diagnoses of ASD (31%) and/or ADHD (31%) were less consistent, autistic features and/or significant behavioral challenges akin to those seen in *RFX3* individuals were reported in the majority of cases, including excitability/overstimulation, sensitivity to sensory (particularly auditory) stimuli, a high pain threshold, emotional dysregulation, aggression, and anxiety (8/8, 100% of those with specific behavioral information provided). Abnormal head size (five individuals with microcephaly and three with macrocephaly) was noted in 8/10 (80%) that provided head circumference measurements. In 4/9 patients (44%) who had neuroimaging, MRI abnormalities were observed (Dandy-Walker malformation, cerebellar tonsillar herniation, an abnormality of the basal ganglia, and a fourth case with limited information but an “abnormal brain MRI” noted). Subtle clinical dysmorphisms were reported in 92% including abnormalities of the hands and feet such as widely spaced toes, syndactyly, or long tapered fingers (50%) (Table 2). Again, no consistent dysmorphisms were evident across individuals.

Five individuals with probable loss-of-function *RFX4* variants were also identified (Tables 1 and 2). Two were individuals who harbored *de novo RFX4* variants, including an in-frame deletion (RFX4 p.(Tyr639_Ser643del)), and a predicted pathogenic missense variant (RFX4 p.(Arg79Ser)) (Table 1, Supplemental Table S2). Three additional related individuals (siblings) were homozygous for a missense variant (p.Thr247Met) altering a well-conserved threonine residue. Of these five individuals, three were female and two were male. Most were noted to have ID (80%) and ASD (80%). All individuals were normocephalic. Neuroimaging was performed in one and demonstrated asymmetric volume loss. Seizures were described in two individuals (40%). No consistent dysmorphisms were evident.

### *RFX3, RFX4*, and *RFX7* variant analyses

In total, 31 distinct variants in *RFX* family members (15 *RFX3*, 3 *RFX4*, and 13 *RFX7* were identified (Table 1). Excluding related individuals, each case involved a novel variant (e.g., there were no recurrent variants). *RFX3, RFX4*, and *RFX7* each exhibit intolerance to loss-of-function variation in human population databases (gnomAD, pLI scores = 1.00). All variants were absent from gnomAD except for *RFX7* p.Pro964_Thr965del, which is detected at a very low frequency in gnomAD v2.1.1 (AF 0.00007677) leading us to formally classify it as a variant of uncertain significance (VUS).

Nineteen protein truncating variants and 12 missense variants were found. We assessed the pathogenicity of the missense variants using six algorithms (MutationTaster, SIFT, PolyPhen2, PROVEAN, LRT, and MutationAssessor) and 11/12 missense variants were predicted to be damaging by at least four algorithms (NsynD score >=4) and the twelfth was predicted damaging by three algorithms (*RFX4* p.(Thr247Met), NsynD3) (Table S2). All missense variants affect highly conserved amino acids (PhastCons vertebrate, mammalian, and primate scores ranging from 0.99-1.00) (Table S2). *RFX* transcription factors are defined by a conserved, specialized winged-helix type DNA binding domain (DBD) that recognize the X-box motif. In addition to the DBD, RFX3 and RFX4 have three known domains that are associated with dimerization (DD).^13^ All variants identified in *RFX3* are located in either the DBD or one of the dimerization domains (15/15, 100%) (Figure 2A, Table 1). *RFX4* and *RFX7* variants did not exhibit clustering to specific functional domains.

We examined 35 additional reported variants in *RFX3, RFX4*, and *RFX7* from prior studies of *de novo* or inherited variants in ASD and neuropsychiatric conditions (Table S3, Figure S1).^4,14,39–42^ Missense variants from the literature tended to be of milder predicted pathogenicity than those reported here (Figure 2C). Sixteen were in *RFX3*, including five *de novo* variants (four protein truncating and one missense variant predicted damaging by all six algorithms, NsynD6, supportive of likely pathogenicity), seven inherited variants (four CNVs, one frameshift variant (p.(Pro408fs)) and three missense variants (p.(Thr151Ala), p.(Ala101Thr), p.(Arg615His); NsynD scores 3-6), and four copy number variants (all microdeletions) were reported for which parental inheritance was not established (Table S3). Among previously reported *RFX7* variants, one was a *de novo* frameshift and one was an inherited frameshift variant. There were also six reported inherited missense variants (6/6 with NsynD >4), and two *de novo* missense variants that are likely benign. Finally, there were nine previously reported *RFX4* variants, only one of which was *de novo* (a missense variant lacking strong evidence of pathogenicity), and eight inherited missense variants of varying predicted pathogenicity (NsynD scores 3-6).

### *RFX* expression is enriched in human brain

*RFX3, RFX4* and *RFX7* have been reported to have relatively high expression in human fetal cortex.^43^ To determine whether specific cell types are affected by *RFX* haploinsufficiency, we examined single-cell transcriptomes from developing and adult human cortex (Figure 3A-F, Figure S2A-B).^32,44^ In developing human cortex, *RFX3* and *RFX7* exhibited the strongest brain expression, with *RFX3* most highly expressed in maturing excitatory upper enriched neurons, *RFX4* most highly expressed in outer radial glia, and *RFX7* most highly expressed in interneurons from the medial ganglionic eminence (Figure 3A-C). We also examined *RFX* expression patterns in the adult human cortex (Figure 3D-F, Figure S2A-B).^32,44^ Again, *RFX3* and *RFX7* exhibited the highest expression. *RFX3* was most highly expressed in glutamatergic layer 2/3 neurons, followed by astrocytes. *RFX7* was expressed in both inhibitory and excitatory neurons. *RFX4* expression was much lower overall, but highest in astrocytes (Figure 3F). These patterns are similar to prior reports from developing and adult mouse neocortex, suggesting that developmental and cell-specific regulation of *RFX* expression is highly conserved during mammalian cortical development.^45^ These expression profiles suggest that *RFX* pathogenic variants may lead to our observed neurodevelopmental phenotypes by altering early developmental cell fates or by impacting the function of upper-layer cortical neurons, astrocytes, and interneurons.

**Figure 3.**
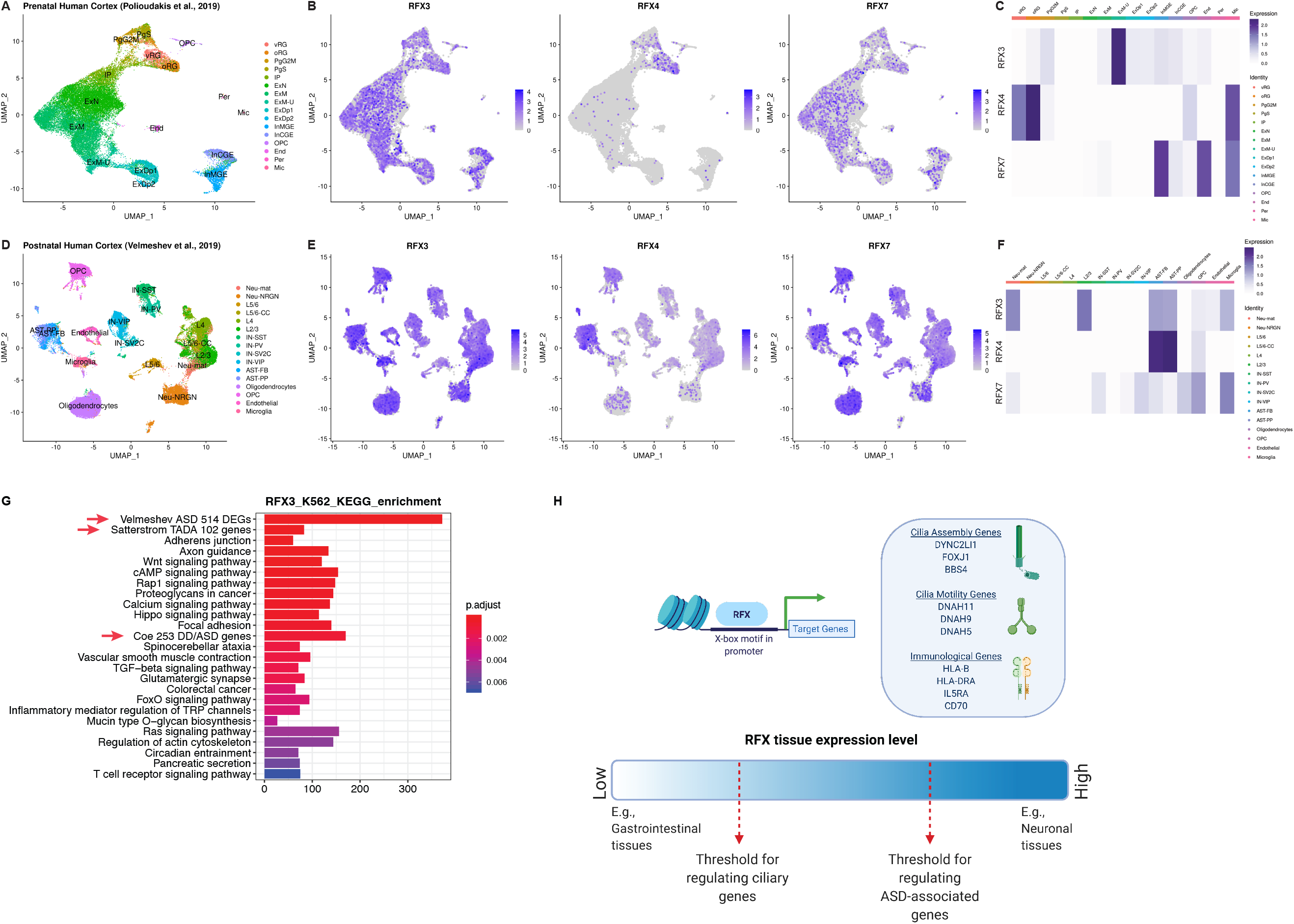
*RFX3, RFX4*, and *RFX7* expression patterns in human cortex and haploinsufficiency gene dosage model. (A) Transcriptomic cell types in the prenatal human cortex identified by single-cell RNA-sequencing.^44^ (B) *RFX3, 4*, and *7* expression patterns in single cells of the prenatal human cortex. (C) Heatmap of *RFX3, 4*, and *7* expression levels among cell types in the prenatal human cortex. (D) Transcriptomic cell types in the postnatal human cortex identified by single-cell RNA-sequencing.^32^ (E) *RFX3, 4*, and *7* expression patterns in single cells of the postnatal human cortex. (F) Heatmap of *RFX3, 4*, and *7* expression levels among cell types in the postnatal human cortex. (G) The enrichment of KEGG pathways, ASD risk gene sets, and ASD differentially expressed genes (DEGs) among RFX3 ChIP-seq binding targets. Pathways and ASD gene sets are ranked by their statistical significance (p.adjust values, Benjamini-Hochberg’s correction). Red arrows indicate ASD risk gene sets and ASD DEGs. X-axis shows the number of genes bound by RFX in their promoter regions. (H) Binding of *RFX* family transcription factors bind to X-box motif in promoter regions of ciliary and immunologic genes. Target gene lists obtained from Durand, Reith, Sugiaman-Trapman.^13,65,66^ Model of *RFX* gene dose-dependent regulation of genes. In tissues with higher expression of *RFX* genes, ASD genes are activated. Lower levels of *RFX* genes are sufficient to activate ciliary genes. vRG, ventricular radial glia. oRG, outer radial glia. PgG2M, cycling progenitors G2/M phase. PgS, cycling progenitors S phase. IP, intermediate progenitors. ExN, migrating excitatory. ExM, maturing excitatory. ExM-U, maturing excitatory upper enriched. ExDp1, excitatory deep layer 1. ExDp2, excitatory deep layer 2. InMGE, interneuron MGE. InCGE, interneuron CGE. OPC, oligodendrocyte precursor cells. End, endothelial. Per, pericyte. Mic, microglia. Neu-mat, immature neurons. Neu-NRGN, NRGN expressing neurons. L5/6, layer 5/6 excitatory neurons. L5/6-CC, layer 5/6 excitatory cortico-cortical projection neurons. L4, layer 4 excitatory neurons. L2/3, layer 2/3 excitatory neurons. IN-SST, somatostatin interneurons. IN-PV, parvalbumin interneurons. IN-SV2C, SVC2 expressing interneurons. IN-VIP, VIP interneurons. AST-FB, fibrous astrocytes. AST-PP, protoplasmic astrocytes. OPC, oligodendrocyte precursor cells.

### *RFX* binding motifs are present in ASD risk gene cis-regulatory regions

Dysregulated gene expression, especially in upper layer cortical neurons, has been implicated in ASD pathogenesis.^32,46^ Given the expression of *RFX3* in layer 2/3 neurons and the autistic features of individuals reported here, we considered whether *RFX* family genes might be important transcriptional regulators of ASD risk genes. RFX family transcription factors bind to a characteristic consensus motif called an X-box (GTHNYY AT RRNAAC)^47^ with individual family members having additional specificity for particular subsequences within this consensus. We therefore performed *RFX3, 4*, and *7* motif enrichment analysis in upstream regulatory sequences of 187 ASD risk genes (the union of 102 TADA genes from Satterstrom et al., 2020 and 124 genes meeting exome-wide significance from Coe et al, 2019) ^14,31^ and an additional set of 447 genes identified (via single cell transcriptome analyses) to be upregulated in ASD brains.^32^ We found enrichment of X-box motifs (q-value <0.05) in human ESC-neuron specific enhancers for ASD risk genes (Table S5A). As a group, *RFX3* and *RFX4* motifs were particularly enriched (q-value <0.005), while the *RFX7*motif was not (q-value 0.48). X-box, *RFX3* and *RFX4* motifs were similarly enriched in the enhancer regions of genes upregulated in ASD brains (Table S5B).^32^ Enrichment of *RFX* motifs in promoter regions of ASD risk genes and DEGs did not emerge (data not shown). Last, we analyzed available RFX ChIP-seq data from the ENCODE project (Table S6) to determine enrichment for Kyoto Encyclopedia of Genes and Genomes (KEGG) pathways, ASD risk gene sets, and ASD DEGs (Table S7). RFX functional binding genes from most ENCODE cell lines were significantly enriched in ASD risk genes and DEGs after multiple testing correction (p.adjust < 0.05; Benjamini-Hochberg’s correction; Figure 3G, Figure S4, Table S8). Across cell lines, there was a positive correlation between enrichment in ASD genes and RFX expression levels in that cell type (Figure S5, Table S9), indicating that higher RFX expression levels may be required to engage ASD relevant targets.

Finally, single gene analyses showed enrichment of *RFX3* and *RFX4* motifs in the promoters of five ASD-associated genes (FIMO p-value <0.0001, q-value <0.1): *AP2S1, KDM6B, ANK2, NONO*, and *MYT1L* (Figure S3B, D),^34^ and *RFX3* ENCODE ChIP-seq data from HepG2 cells confirmed RFX3 binding peaks in the promoters of AP2S1, KDM6B, and NONO (Figure S3E-G). Notably, *de novo* loss-of-function variants in *KDM6B* cause a neurodevelopmental syndrome that has phenotypic overlap with *RFX3* haploinsufficiency as described in this report, namely mild global delays, delayed speech, hypotonia, and features of ASD and ADHD, while loss-of-function variants in *NONO* and *MYT1L* are a cause of X-linked and autosomal dominant intellectual disability, respectively (Table S4).^48–55^ These cases support the model that *RFX* members may be transcriptional activators of a subset of ASD risk genes via actions at both enhancer and promoter sites.

## DISCUSSION

Our results delineate a novel human neurobehavioral syndrome including ASD, ID and/or ADHD due to deleterious variants in *RFX* family transcription factors. While presence of neuroimaging findings, seizures, and dysmorphisms varied between different *RFX* family members, the behavioral phenotypes of individuals with *RFX3, RFX4*, and *RFX7* were strikingly similar, and often included sensory hypersensitivity and impulsivity.

This report complements accumulating statistical genetic evidence for *RFX3* as an ASD risk gene,^14,41^ and extends these findings to the closely related *RFX* family members *RFX4* and *RFX7*. Two-thirds of individuals with *RFX3* variants in our series carried an ASD diagnosis, half had ADHD, and just over half of individuals had ID. Several individuals with *RFX3* variants also exhibited post-pubertal cognitive or behavioral regression sometimes accompanied by psychosis. *RFX3* CNVs have been previously reported in schizophrenia^40,56^

In contrast to individuals with *RFX3*, individuals with *RFX7* variants showed almost uniform diagnoses of language delay and ID (92%). Individuals with *RFX7* variants were less likely to have ASD or ADHD diagnoses. Fewer individuals were identified with *RFX4* variants, but those identified had high rates of ASD and ID.

*RFX* family members have been previously known for their biological roles in cilia development. The *RFX3* transcription factor activates core components necessary for development and maintenance of both motile and immotile cilia,^57–59^ and biallelic *Rfx3* knockout in mice results in situs inversus, hydrocephalus, and deficits in corpus callosum formation.^59–61^ This raises the question of whether the neurodevelopmental phenotypes reported here may be mechanistically related to cilia development – e.g., a hypomorphic human ciliopathy. The majority of described genetic ciliopathies are recessive, and therefore not due to haploinsufficiency, but some (e.g. Meckel syndrome, Joubert syndrome, Bardet-Biedl syndrome, oral-facial-digital syndrome type I) may be associated with neurodevelopmental abnormalities and/or brain malformations. Conversely, the individuals described in this report lack systemic features of ciliopathies.

In a competing model, the neurodevelopmental consequences of *RFX* haploinsufficiency may reflect non-ciliary roles of RFX transcription factors in brain development (Figure 3H). Finally, while not formally included in the case series presented here, we make note of an *RFX4* case that was ascertained after an early pregnancy termination and noted to have bilateral ventriculomegaly on autopsy. The fetus was determined to be a compound heterozygote for missense variants in *RFX4* that are of uncertain significance (maternally inherited c.55C>T, p.(Arg19Trp) and paternally inherited c.1519A>T, p.(Ile507Phe)), raising the possibility that biallelic mutations in *RFX* genes may lead to more severe phenotypes than those described in this series.

Enriched expression of *RFX3* in upper cortical layer neurons places this gene in cells that are involved in communication between regions of the cortex important for higher cognition and social behavior,^62^ raising the possibility that haploinsufficiency may disrupt either the developmental specification, synaptic connectivity, or electrophysiological function of this set of neurons. Projection neurons in this layer have been implicated in ASD by analyses of co-expression networks of autism genes,^63,64^ and superficial cortical neurons exhibit the strongest amount of differential gene expression in ASD brains compared to controls.^32,46^ Sun and colleagues in fact showed strong enrichment of RFX motifs in differentially acetylated peaks upregulated in ASD brains compared to controls.^46^ Future studies aimed at understanding the downstream targets of RFX family members in human brain may shed new light on pathways important to the molecular pathogenesis of ASD, ADHD, and ID.

## Data Availability

All data referred to in this manuscript is either provided in the main text/ supplementary material, or appropriately referenced (where derived from pre-existing, publicly accessible datasets).

## SUPPLEMENTAL DATA

Supplemental Data include detailed clinical descriptions for each affected individual, five figures, and nine tables.

## ACKNOWLEDGEMENTS

We thank the patients and their families for their participation and inclusion in this work. We also thank all the genetic counselors working with our patients and families. We thank the DDD study for providing exome sequencing results. We thank Billie Lianoglou, MS, CGC (Fetal Treatment Center, University of California at San Francisco Fetal Treatment Center, San Francisco, CA, 94158, USA) for her contributions of an RFX4 variant of unclear significance (see supplementary material). We thank GeneDx for their contributions as a collaborator via GeneMatcher, and Ambry Genetics for their contributions including facilitation of clinician communication. JL was supported by award T32GM007753 from the National Institute of General Medical Sciences. The content is solely the responsibility of the authors and does not necessarily represent the official views of the National Institute of General Medical Sciences or the National Institutes of Health. BZ was supported by the Manton Center Pilot Project Award and Rare Disease Research Fellowship. JEP and JRL were supported in part by the National Human Genome Research Institute (NHGRI) and National Heart Lung and Blood Institute (NHBLI) to the Baylor-Hopkins Center for Mendelian Genomics (BHCMG, UM1 HG006542). KU, SAB, CF, and SR were supported by the French Ministry of Health and the Health Regional Agency from Bretagne, Pays de la Loire and Centre Val de Loire (HUGODIMS 2, 2017). The DDD study is supported by the Wellcome Trust and the UK Department of Health Innovation Challenge Fund [HICF-1009-003] and the Wellcome Trust Sanger Institute [grant no. WT098051] (Nature 2015;519:223-8). TWY was supported by grant nos. NIH/NIMH R01MH113761, NICHD/NHGRI/NIH U19HD077671 and NIH/NICHD U24HD0938487, and by a SFARI Pilot Research Award.

## AUTHOR CONTRIBUTIONS

TWY, HKH, and TN conceptualized the paper, collected case information from collaborators, conducted and managed all functional studies, drafted the initial manuscript, and edited and revised the manuscript. TWY provided research study oversight. JL designed and performed *RFX* motif analysis, analyzed published brain transcriptome and single-cell RNA-sequencing data, analyzed *RFX3, 4*, and 7 variants and their predicted pathogenicities, cataloged *RFX* variants in previously published studies and analyzed their pathogenicities, contributed to the manuscript, and edited and revised the manuscript. BZ collected ChIP-seq data and performed over-representation analysis, contributed to the manuscript, and edited and revised the manuscript. NA and CSG conducted functional studies of the impact of *RFX3* variants on protein stability. AS performed RFX variant analyses. CAG assisted in research enrollment. LR, RP, TG, BBAdV, MEHS, KLIvG, EvB, C(N)MLV, AH, CDA, LLI, CB, MW, EF, TLT, KWG, LB, FV, PR, XW, JLA, MF, GET, JEP, JRL, EA, AN, RA, ARa, PB, CRF, MJL, MK, GL, AL, AP, KKP, LEW, KA, JB, CS, JM, CPB, GP, PG, MB, SK, MN, IGR, MYZ, CK, ARe, MI, KU, SA, CF, SR, MI, PDT, JB, YW, GZ, SS, IB, RAJ, WBD, and LLC contributed clinical case information and/or analyzed exome data. PBA and AB supported research subject enrollment.

## SUPPLEMENTAL FIGURE TITLES AND LEGENDS

**Figure S1. Pathogenicity scores of missense variants in *RFX3, RFX4*, and *RFX7***.

(A) The *RFX3* missense variants reported in this study have a higher frequency of pathogenic predictions than prior reported missense variants.

(B) *De novo* missense variants are more likely pathogenic than inherited missense variants.

(C) *RFX3* missense variants are more likely damaging than *RFX4* or *7* missense variants.

(D) Prior reported missense variants in *RFX3, 4*, and *7* do not show clear associations between predicted pathogenicity and inheritance.

MPC, Missense badness, PolyPhen-2, and Constraint. NsynD, Nonsynonymous Damaging score. CADD, Combined Annotation Dependent Depletion.

**Figure S2. *RFX3, RFX4*, and *RFX7* expression patterns in human cortex**.

(A) *RFX3, 4*, and *7* expression patterns in single cells of adult human cortex (Allen Human Brain Atlas).^67^ Cell types identified in human cortex shown in t-SNE plot with taxonomy of clusters. Expression color-scale units are Log2(CPM+1). Data from Allen Brain Map single-nucleus RNA-sequencing of human cortex (Image credit: Allen Institute).

(B) Heatmap visualizing *RFX3, 4*, and *7* expression among adult human cortical cell types, along with canonical cell type markers. (Image credit: Allen Institute).

(C) *RFX3, RFX4*, and *RFX7* expression across all human tissues. (Data Source: GTEx Analysis Release V8 dbGaP Accession phs000424.v8.p2).

**Figure S3. Presence of RFX3 and RFX4 binding motifs in ASD-associated gene promoters**.

(A) RFX3 binding motif MA0798.1 (JASPAR 2020).

(B) ASD risk genes with significant RFX3 binding motif occurrences.

(C) RFX4 binding motif MA0799.1 (JASPAR 2020).

(D) ASD risk genes with significant RFX4 binding motif occurrences.

(E) – (G) RFX3 ChIP-seq binding peaks are located in promoter region of ASD-associated genes (E) AP2S1, (F) KDM6B, (G) NONO.

All motif occurrences were identified using FIMO, q-values <0.10. RFX3 in HepG2 ChIP-seq binding peaks obtained from ENCODE GSM2534235.

**Figure S4. Customized KEGG enrichment analysis of RFX functional binding sites**.

The enrichment of KEGG pathways, ASD risk gene sets, and ASD differentially expressed genes (DEGs) for different RFX binding profiles. (A–C) representative barplots of the enriched customized KEGG pathways were shown using RFX1 (A), and RFX5 (B-C) ChIP-seq data. Pathways and ASD gene sets are ranked by their statistical significance (p.adjust values, Benjamini-Hochberg’s correction). Red arrows indicate ASD risk gene sets and ASD DEGs. X-axis shows the number of genes bound by RFX in their promoter regions. Data used for Figure S4D were obtained from the GTEx Portal, dbGaP accession number phs000424.v8.p2 on 05/01/2020.

**Figure S5. Correlation of enrichment significance in ASD gene sets with *RFX* expression level**.

*RFX* expression levels in different cell lines were correlated with the enrichment significances [–log10(p.adjust)] for (A) ASD 514 DEGs; (B) TADA 102 ASD risk genes; (C) 253 ASD/ID risk genes. Red dotted line indicates the P-value = 0.05. X-axis shows the GTEx expression of *RFX* in different cell lines derived from a variety of human tissues.

## SUPPLEMENTAL TABLE TITLES AND LEGENDS

**Table S1. Summary of clinical features associated with pathogenic variants in *RFX3, RFX4* or *RFX7***.

Clinical features summarized by the number of positive reports out of the total number of individuals per gene, with the exception of “Micro or Macrocephaly” and “Neuroimaging findings,” where the fraction is the number of positive reports out of the total number of reports available per gene. Proportions are shown in parentheses.

**Table S2. Predicted pathogenicity of *RFX* variants**.

NA, not applicable. Italics indicates individual has a variant of uncertain significance.

**Table S3. *RFX* variants referenced in prior published studies of neurodevelopmental delay**.

RefSeq identifiers: RFX3 NM_134428.2, NP_602304.1; RFX4 NM_213594.2, NP_998759.1; RFX7 NM_022841.5, NP_073752.5. gDNA coordinates are provided as originally reported by each reference (Hg19: Krumm et al., 2015, Li et al., 2017, Sahoo et al., 2011, Walsh et al., 2008. Hg38: De Rubis et al., 2014, Tabet et al., 2015). AD, activating domain. DBD, DNA binding domain. DD, dimerization domain. NA, not applicable.

**Table S4. Phenotypes associated with ASD risk genes potentially regulated by *RFX* members**.

De novo loss of function variants in *KDM6B, NONO, and MYT1L* are associated with NDD/ID phenotypes. These phenotypes are similar to those seen in the individuals with *RFX* variants in this present study. AD, autosomal dominant. XL, X-linked.

**Table S5. Enrichment of RFX binding motifs in enhancer regions associated with ASD risk genes and ASD upregulated genes in human embryonic stem cell (ESC)-derived neurons**.

A) RFX binding motifs enriched in human ESC-neuron enhancer regions associated with ASD risk genes, q-value <0.05. B) RFX binding motifs enriched in human ESC-neuron enhancer regions associated with ASD upregulated genes, q-value <0.05. Lack of enrichment for RFX binding motifs in promoters of ASD upregulated genes. Fold-enrichment reported as the % motifs identified in target sequences/% motifs identified in background sequences. All motif enrichment analyses were performed using HOMER tool.

**Table S6. ChIP-seq and eCLIP datasets from the ENCODE project**.

**Table S7. Customized ASD risk and ASD DEG gene sets**.

**Table S8. Enrichment statistics of customized KEGG analysis**.

**Table S9. Correlation between RFX expression level and enrichment for ASD gene sets**.

